# Improved biorepository to support sickle cell disease genomics and clinical research: A practical approach to link patient data and biospecimens from Muhimbili Sickle Cell Program, Tanzania

**DOI:** 10.1101/2023.01.06.23284272

**Authors:** Upendo Masamu, Raphael Z Sangeda, Josephine Mgaya, Siana Nkya, Beatrice Octavian, Frank R Mtiiye, Joyce Nduguru, Agnes Jonathan, Daniel Kandonga, Irene K Minja, Paschal Rugajo, Emmanuel Balandya, Julie Makani

## Abstract

Genetic modifiers underlying various sickle cell disease phenotypic expressions are largely unexplored in Africa due to lack of databases linking biospecimens with demographic and clinical data. The problem may be compounded by a complete lack of biorepositories in these settings. This article aims to document the physical verification of biospecimens stored in the biorepository and link them to patient clinical and demographic information to facilitate plans for genomic and related clinical research studies. We reviewed and updated the existing biorepository infrastructure at Muhimbili Sickle Cell Programme in Dar es Salaam, Tanzania. The database of archived biospecimens was updated with the location information of respective biospecimens following the physical verification of biospecimens and then mapping the patient demographic and clinical data with the biospecimen data using sickle cell patients’ demographic identifiers. Three freezers maintained at -80°C store a total of 74,079 biospecimens, of which 63,345 were from 5,159 patients registered in the Muhimbili Sickle Cohort from 2004 to 2016. Out of stored biospecimens, follow-ups were 46,915 (74.06%), control 8,067 (12.74%), admission 5,517 (8.71%) and entry 2,846 (4.49%). Of these registered patients, females were 2,521 (48.87%) and males were 2,638 (51.13%). The age distribution was 1 to 59 years, with those above 18 years being 577 (11.18%) and children 4,582 (88.82%) of registered patients. The notable findings during the process include a lack of automated biospecimen checks, laboratory information management system and standardization of equipment used, biospecimens not linked to clinical and demographic data, date format inconsistencies, lack of regular updating of a database on exhausted biospecimens and updates when biospecimens are moved between positions within freezers. Well-organized biorepository plays a crucial role in answering future research questions. Enforcing strict standard operating procedures and quality control standards will ensure that laboratory scientists and other users adhere to the best biospecimen management procedures.

## Introduction

Sickle cell disease (SCD) is an inherited disease caused by a single-gene mutation affecting the β-globin gene on chromosome 11. It results in an abnormal hemoglobin protein (HbS) which affects the shape and function of red blood cells (RBC) and subsequently impacts nearly all body organ systems [1,2]. SCD is one of the most prevalent inherited blood disorders. An estimated 300,000 individuals are born each year with SCD, the highest burden being in Africa, where up to 75% of SCD births occur [3]. It is estimated that 50–80% of infants born with SCD in Africa die before the age of 5 years [4]. Research on SCD is crucial, allowing the discovery of a cure that will reduce its burden on afflicted families. Therefore, it is imperative that the current and future research on SCD store and maintain biospecimens appropriately linked to their clinical and demographic information.

Biorepositories hold a crucial role in achieving biomedical progress and improving research capacity in Africa [5–7], given the advancements in technology in medical sciences. Biospecimens should be stored in a manner to support not only current use but also future clinical or research studies. Human biorepositories are used to understand diseases and diagnoses and ultimately provide information on the cure and treatment [8,9]. Several initiatives have been established to support African biorepository activities. Such initiatives include the Human, Heredity, and Health in Africa (H3Africa), which has helped several biorepositories such as the Integrated Biorepository at Makerere University College of Health Sciences (IBRH3AU), Institute of Human Virology (IHV) H3Africa Biorepository in Abuja Nigeria Clinical Laboratory Services (a Division of the Wits Health Consortium) in Johannesburg South Africa. According to the literature, Gambia was the first country in Africa to establish a national DNA bank [10,11]. Sustainability and the ability to answer research questions are critical for a standard biorepository [12–15]. A well-maintained biorepository is expected to have stored biospecimens to ensure quick biospecimen retrieval and the validity of the analytical tests. This includes a clear link between the biospecimen data and clinical and demographic information, increasing the scientific value of stored biospecimens [16,17]. This can be achieved if clearly defined procedures govern biorepository processes. Several discussions have been around regarding the standards that any biorepository is supposed to have. One of the discussions is on standard operating procedures (SOPs) for biorepository processes such as sample collection, storage, processing, and information management such as Laboratory information management system (LIMS) [16,18,19]. There are few established biorepositories for SCD globally. These establishments would speed the discovery of mitigations against SCD, including discovering novel biological markers for treatment.

The Muhimbili Sickle Cell Programme at the Muhimbili University of Health and Allied Sciences (MUHAS) in Tanzania was established in 2004 and aimed to offer clinical care and build research capacity in SCD [1,3]. The program recruited a Muhimbili Sickle Cell Cohort (MSC) database of over 5,000 SS individuals, including their demographic, clinical, and biospecimens data. The main goal of the program was to conduct research on SCD that will identify the disease spectrum, causes of disease morbidity and mortality, and describe genetic determinants of the disease [20–25]. Following the research impact achieved with the MSC[3], MUHAS was awarded twice a grant named Sickle Pan African Consortium (SPARCo) by the National Institutes of Health (NIH). This was a collaborative grant between MUHAS in Tanzania as a research hub and a site, Cape Town University as a data coordinating center and two other research sites in Nigeria and Ghana in phase 1 between 2017 and 2021. In phase 2, granted between 2021 and 2026, additional three sites in four countries (Mali, Uganda and a shared site in Zimbabwe and Zambia) were established. SPARCo plans to conduct genomic and clinical studies in the second phase of the SPARCo grant award [26].

The challenge in Muhimbili Sickle Program and other centers in African counties is the ability to manage and trace biospecimens that move between different positions and locations in the biorepository. In addition, linking the biospecimen data to the clinical and demographic data is tedious without an automated way of handling and storing biospecimens. Linking the biospecimens with their clinical and demographic data is essential because it may give insight into the pattern that may accelerate SCD research progress and discovery. Furthermore, automatic handling and storing of biospecimens is necessary to allow biospecimen integrity and reproducibility of results [19,27]. To achieve this goal, this study was designed to facilitate genomics research by preparing easily maintainable records of biospecimens from the previous MSC cohort and new biospecimens from patients. We describe the experience of conducting quality control and verifying all archived biospecimens from Muhimbili Sickle Program and Sickle Pan African Research Consortium (SPARCo) and integrate the biospecimen database with the clinical and demographic data using the Research Electronic Data Capture (REDCap). The system established will allow quick and intuitive data management that is scalable. The documentation of quality assured biorepository shall serve as the basis for other SSA countries to emulate in practice and pave the way for collaborative genomic studies utilizing the existing biospecimens.

## MATERIALS AND METHODS

### Study design and population

The study was conducted among patients who consented to participate in the MSC study and its nested studies, such as the Vascular Function Intervention Trial (VFIT) and Strategic Award Study [3,28]. The enrollment into the MSC included all individuals who attended a clinic or patients suspected clinically diagnosed to have SCD at Muhimbili National Hospital (MNH) in Dar es Salaam, Tanzania. Patients were registered mainly from the coastal regions and other regions in Tanzania.

### Ethics Statement

The Senate Research and Publications Committee of the Muhimbili University of Health and Allied Sciences (MUHAS) in Tanzania approved the study (Ref. No.DA.282/298/01.C/). Written informed consent was obtained from all participants. For individuals 18 years or above, while for minors, a parent/ guardian consented and signed the consent on behalf of the patient; adolescents provided assent.

### Biospecimens verification

The study reviewed, quality assured and updated the existing biorepository infrastructure at the program to link SCD biospecimen data with their clinical and demographic data. During the MSC, patients were identified at entry, admission, follow-up, and as a control which were recorded as visit types. Different biospecimens were collected, including plasma, buffy coat, serum, DNA and isolates. These biospecimens were stored in microtubes within a cryobox, whereby one cryobox would only store one biospecimen type for a specific study. The cryoboxes were labeled on top with the study name, biospecimen type and box number. The numbers on the cryobox were unique for each biospecimen type throughout the study. Basic clinical and demographic information was collected using case report forms (CRF).

Physical verification of biospecimen location within a freezer was done by verifying information such as freezer number, a compartment within a freezer, rack and box number. Information regarding the biospecimen, such as type and study, were also verified during the process. Data from physical verification were compared to the one on the database and updated corresponding records. The final database was cleaned up by maintaining all records of biospecimens found in the freezers during physical verification and having all the verified information about the biospecimens and freezer locations.

Tubes found empty and still in the freezers were removed and updated in the database. The clean biospecimen data was mapped and linked to its clinical and demographic information using demographic identifiers through which two categories were identified cohort-related for whom demographic identifiers were available and non-cohort for biospecimens from patients not admitted in the MSC cohort. The clean final biospecimen data were migrated to the REDCap database to allow easy record-keeping and retrieval. A REDCap (Research Electronic Data Capture) database was created to store curated biospecimens and patient data from this study. REDCap is a secure, web-based application designed to support data capture for research studies, providing (1) an intuitive interface for validated data entry; (2) audit trails for tracking data manipulation and export procedures; (3) automated export procedures for seamless data downloads to standard statistical packages; and (4) procedures for importing data from external sources [29,30]. The REDCap system and tools were hosted at Muhimbili University of Health and Allied Science.

### Data analysis plan

Descriptive such as counts, averages and proportions were used to summarize patients’ demographic and biospecimen data. Rstudio 2022.07.2 was used for summarizing the data.

## Results

The Hematology Clinical and Research Laboratory (HCRL) at Muhimbili has three freezers (Fig1A) used to store the biospecimens from MSC. The freezer has three areas within a compartment, columns, and rows (Fig1B). Biospecimens of types of plasma, buffy coat, deoxyribonucleic acid (DNA), serum, urine, washed red blood cells and bacterial isolates were aliquoted in the 0.5-2uL microtubes and arranged in 9×9 cryoboxes. The freezers were monitored for temperature checks twice daily and adequately documented. A backup generator is connected to the main power of the laboratory to observe the quality of the archived biospecimens in case of a power failure. Currently, the HCRL contains a total of 74,079 biospecimens. The space occupied by these biospecimens was 70% of the available space for archived biospecimens stored in the three -80°C freezers.

**Fig 1:**
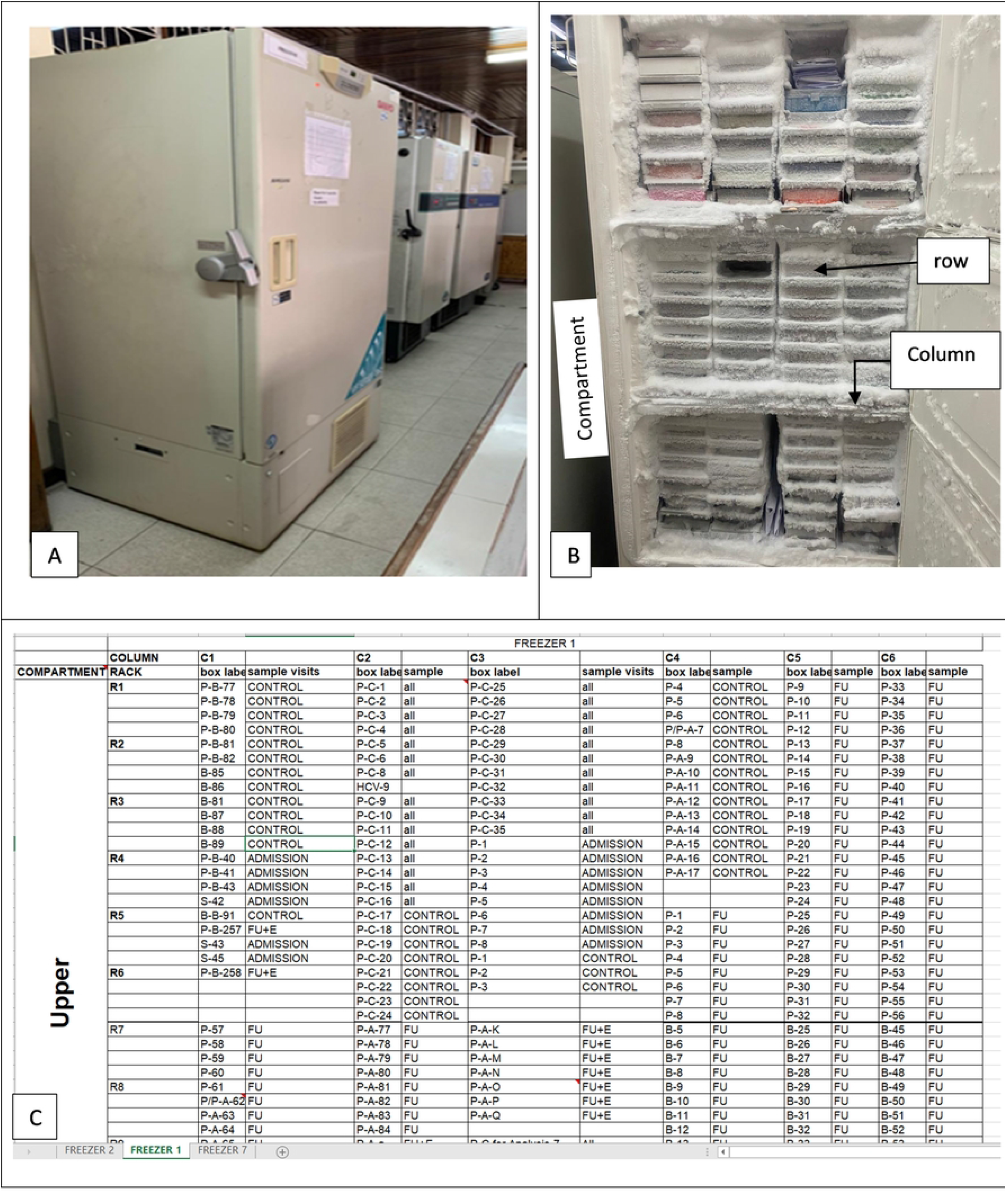
Freezers with the archived biospecimens. (A) -80C freezers for storing archived biospecimen. (B) A picture showing the inside of one of the freezers. (C) Freezer plan map showing cryobox positions within the freezer (Compartment, C-Column and R-Rack, Biospecimen type: P-Plasma, FU-Follow-up, E-Entry, S-Serum)

Out of 74,079 archived biospecimens, 63,345 biospecimens were from 5,159 patients registered under MSC. The remaining 10,734 biospecimens were from nested studies within the MSC. More patients were seen at the inception of the project in 2004, but in 2016 new clinics were opened at other referral hospitals that patients were referred to instead of coming to Muhimbili (Fig 2). Of 5,159 registered patients, the females were 2,521 (48.87%), while the males were 2,638 (51.13%). In terms of age distribution, the minimum age was less than one year while the maximum age was 59, with those above 18 years being 577 (11.18%), while children were 4,582 (88.82%) of registered patients under MSC ‘Table 1’. The total number of biospecimens collected per patient is categorized in Fig 3. The group labeled 1 is the group with patients with only one biospecimen collected and others are in intervals of 10 biospecimens.

**Fig 2:**
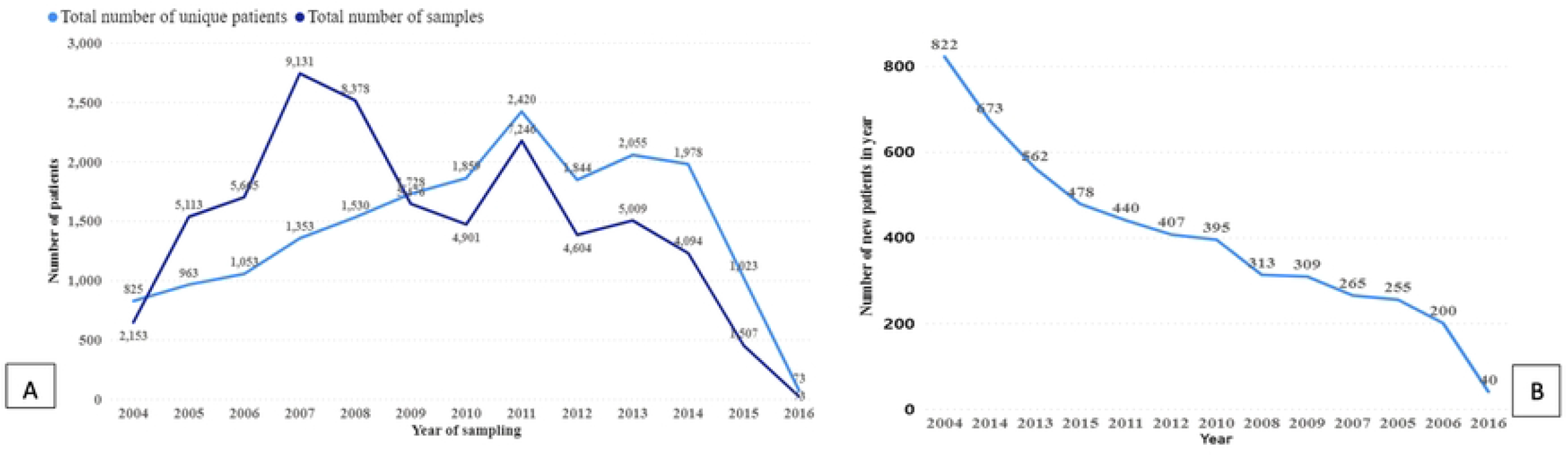
Patients enrollment trend and biospecimen collection (A) Number of unique visiting patients and biospecimens collected over the years 2004-2016 in Muhimbili Sickle Cohort (MSC); (B) indicates the number of new patients enrolled each year.

**Table 1.**
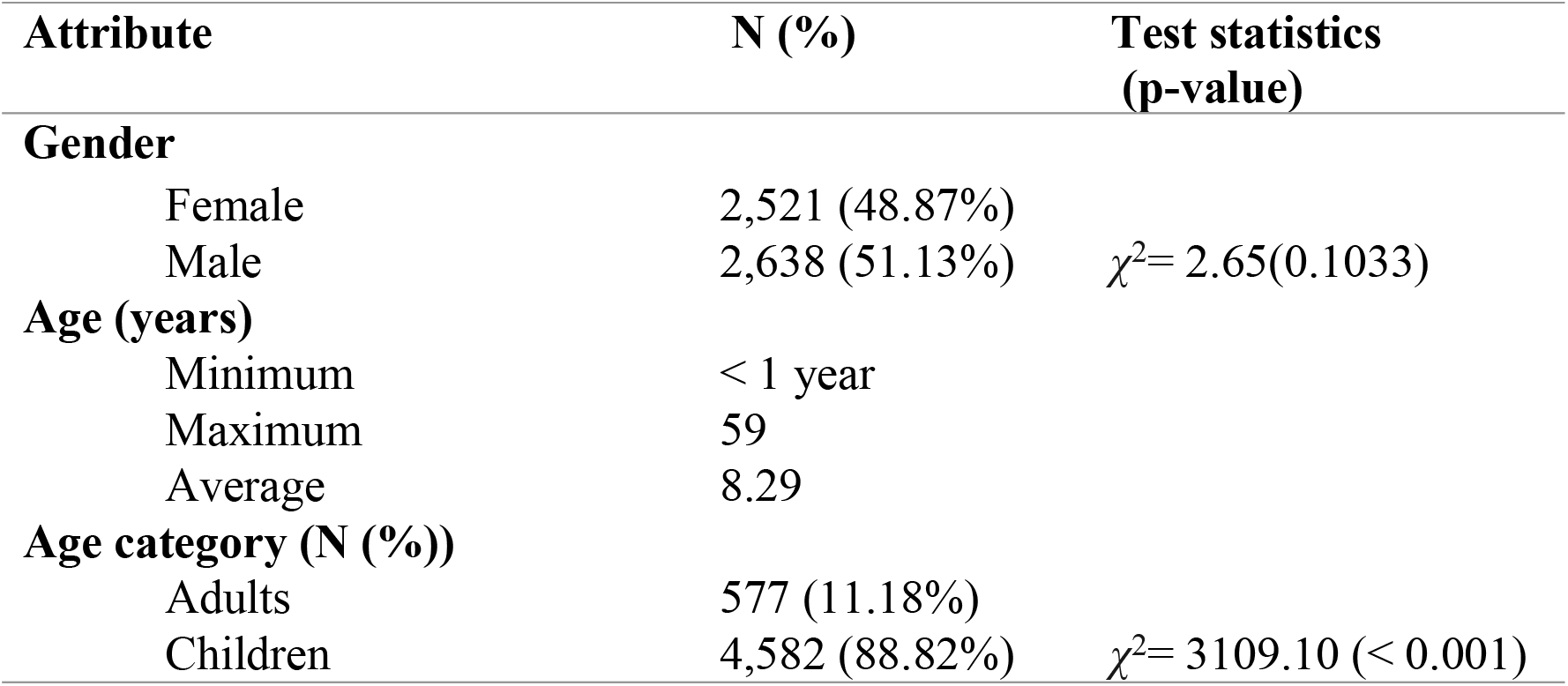
Demographic characteristics of patients under the Muhimbili Sickle Cohort

**Fig 3:**
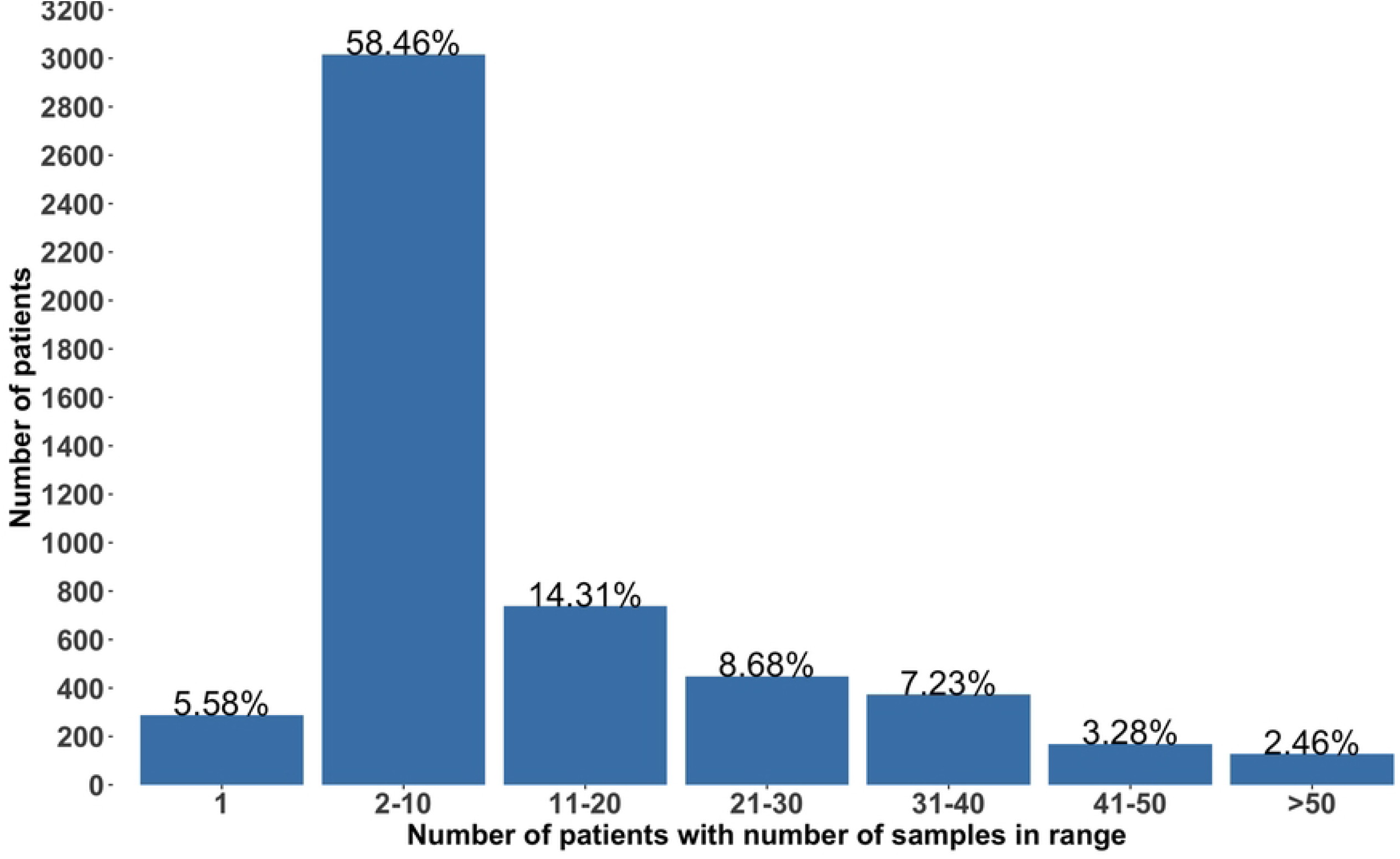
Number of patients with a given number of biospecimens stored in the biorepository.. The group labeled 1 is the group with patients with only one biospecimen collected and others are in intervals of 10 biospecimens.

Out of 63,345 biospecimens that were collected per each visit type follow-ups were 46,915 (74.06%), control 8,067 (12.74%), admission 5,517 (8.71%) and entry 2,846 (4.49%) ‘Table 2’. From the physical verification, biospecimen types present include plasma 38,008 (60.00%), buffy coat 15,323 (24.19%), serum 4,671 (7.37%), DNA 4,528 (7.15%) and bacterial isolates 258 (0.41%) ‘Table 3’. The 557 biospecimens ‘Table 3’ were from a nested study on Amino Acids in Tanzanian Children with Sickle Cell Disease under Vascular Function Intervention Trial (V-FIT)[31]. One of the outcomes of the verification process was to generate and update freezer plan maps, usually placed in front of the freezer showing the position of boxes with biospecimens within the freezers. The map captures the biospecimen type, box number, compartment, rack position within a freezer and the visit type (Fig 1C).

**Table 2.** Number of archived biospecimens per year and visit type under Muhimbili Sickle Cohort

| Year | Admission | Control | Entry | Follow-up | Year total |
| --- | --- | --- | --- | --- | --- |
| 2004 | 34 | 10 | 965 | 1,144 | 2,153 (3.40%) |
| 2005 | 442 | 154 | 255 | 4,261 | 5,112 (8.07%) |
| 2006 | 224 | 275 | 381 | 4,785 | 5,665 (8.94%) |
| 2007 | 359 | 859 | 336 | 7,575 | 9,129 (14.41%) |
| 2008 | 619 | 1,221 | 266 | 6,270 | 8,376 (13.22%) |
| 2009 | 564 | 423 | 80 | 4,409 | 5,476 (8.64%) |
| 2010 | 529 | 462 | 63 | 3,847 | 4,901 (7.74%) |
| 2011 | 857 | 1,025 | 106 | 5,258 | 7,246 (11.44%) |
| 2012 | 607 | 696 | 139 | 3,162 | 4,604 (7.27%) |
| 2013 | 568 | 1,032 | 154 | 3,255 | 5,009 (7.91%) |
| 2014 | 391 | 1,232 | 78 | 2,393 | 4,094 (6.46%) |
| 2015 | 323 | 637 | 23 | 524 | 1,507 (2.38%) |
| 2016 |  | 41 |  | 32 | 73 (0.12%) |
| **Visit type total** | **5,517**<br>**(8.71%)** | **8,067**<br>**(12.74%)** | **2,846**<br>**(4.49%)** | **46,915**<br>**(74.06%)** | **63,345** |

**Table 3.** Number of each archived biospecimen type under the Muhimbili Sickle Cohort

| Year | Buffy coat | DNA | Isolates | Plasma | Serum | V-FIT |
| --- | --- | --- | --- | --- | --- | --- |
| 2004 | 589 | 20 |  | 1,153 | 391 |  |
| 2005 | 2,407 | 93 |  | 2,420 | 192 |  |
| 2006 | 2,121 | 240 | 69 | 2,933 | 302 |  |
| 2007 | 1,449 | 1,829 | 37 | 4,699 | 1,115 |  |
| 2008 | 738 | 673 | 95 | 6,292 | 578 |  |
| 2009 | 603 | 205 | 54 | 3,857 | 757 |  |
| 2010 | 254 | 296 | 3 | 4,139 | 209 |  |
| 2011 | 1,339 | 1,172 |  | 4,365 | 370 |  |
| 2012 | 1,624 |  |  | 2,559 | 253 | 168 |
| 2013 | 1,993 |  |  | 2,487 | 223 | 306 |
| 2014 | 1,604 |  |  | 2,267 | 140 | 83 |
| 2015 | 550 |  |  | 816 | 141 |  |
| 2016 | 52 |  |  | 21 |  |  |
| **Biospecimen type total** | **15,323**<br>**(24.19%)** | **4,528**<br>**(7.15%)** | **258**<br>**(0.41%)** | **38,008**<br>**(60.00%)** | **4,671**<br>**(7.37%)** | **557**<br>**(0.88%)** |

## Discussion

This paper documents best practices and challenges from verifying archived biospecimens and experience at the Muhimbili Sickle Cell Program in Dar es Salaam, Tanzania. Physical verification of archived biospecimens was carried out whereby a total of 74,079 biospecimens were archived, of which 63,345 biospecimens were from 5,159 patients registered under MSC. Different biospecimens, such as buffy coat, plasma, DNA, serum and bacterial isolates, were archived.

During the physical verification process, some observations needed to be addressed to have a sustainable biorepository to answer future research questions. The absence of an automated way of handling biospecimens made the verification process tedious and manual. The lack of automatic biospecimen checks added to the long, tedious biospecimen verification process. Institutions planning to have biorepositories should consider having an automated way of handling biospecimens, such as a Laboratory Management Information System (LIMS), to allow automatic biospecimen checks that will enable fast and efficient tracking of archived biospecimens. The use of an automated way of handling and storing biospecimens was also recommended by other researchers to preserve biospecimen integrity and allow the reproducibility of results [19,27,32,33].

Some of the tubes with archived biospecimens were missing date labels that indicate when the biospecimens were taken. The recording of the date on the tube needs to be enforced because biospecimens are only kept for a specific period and those with no dates may need to be discarded. Proper date annotation is critical to allow biospecimens’ data integrity and quality [17,19]. In addition, some of the archived biospecimens were exhausted in the freezer, but their information was not updated in the database. This calls for laboratory scientists to regularly update the database with information on the exhausted biospecimens. This will allow other users to be informed of the finished biospecimens. The process can be enforced and make laboratory scientists more accountable by implementing a LIMS that tracks users’ activities by their identifiers [32].

Lack of standards on the type of tube to be used, whereby some tubes had volume labels while some did not. The absence of a volume label caused difficulties verifying the volume for the archived biospecimens. This calls for developing an SOP on the standard of the types of equipment to be used for biospecimen archiving, which will guide the procurement of respective equipment. Other researchers also recommended the need to have SOPs for the procurement of biospecimen archiving equipment [8].

Different recorded date formats from multiple users who entered data in Microsoft Excel. The date format challenge can also occur when a proper database system is in place. However, the database management engine usually handles date format issues more conveniently. Internet connectivity is another challenge in Africa, preventing real-time access to LIMS. In such cases, laboratory scientists may use MS Excel to temporarily log data and then later import it to an internet database via LIMS. Hence to overcome these challenges, there is a need to have an SOP in place that map the process of recording information for the biospecimen [8]. Such SOP includes a clause defining a date format to be used and adhered to.

It was also observed that some biospecimens were moved to new positions within a freezer, but that information was not updated in the biospecimens’ database. This calls for logging to track biospecimens’ movement within the freezer to account for biospecimen misplacement. Furthermore, a few tubes were empty and still found in the freezer. Strict rules and SOP should be implemented to guide laboratory scientists on biospecimen management. Such rules include removing empty tubes once exhausted and updating the database. In addition, there should be ownership of the biospecimen management systems among the users. This means a person who oversees and enforces all the rules should be answerable to all queries [34]. Staff training and periodic review of processes are essential to ensure that SOPs are being adhered to [33,34].

## Conclusion

The biorepository plays a crucial role in answering future research questions to allow the discovery of novel genomic markers for diagnosing and treating SCD. However, this will be achieved if biospecimens are adequately managed. Strict controlled standard operating procedures and quality assured and quality controlled standards must be enforced to ensure that laboratory scientists and other users adhere to the best biospecimen management procedures. This includes SOP for the procurement of standard equipment, biospecimen storage (such as biospecimen labeling during archiving), and movement from and to the freezers. Regular training on best practices and SOPs for new staff and refresher training for existing ones. Furthermore, one should have LIMS and automatic checks that will allow fast and efficient tracking of archived biospecimens.

## Data Availability

The data of this study are available from the corresponding author at a reasonable request.

## Acknowledgments

We would like to thank SCD patients whose data were used in this study and all staff at the Muhimbili Sickle Cell Program for their support.

## Author Contributions

**Conceptualization:** Raphael Z Sangeda

**Investigation:** Upendo Masamu, Josephine Mgaya, Beatrice Octavian and Frank R Mtiiye

**Formal analysis and Visualization:** Upendo Masamu and Raphael Z Sangeda

**Writing – Original Draft Preparation:** Upendo Masamu, Raphael Z Sangeda, Josephine Mgaya, Siana Nkya, Beatrice Octavian, Frank R Mtiiye, Joyce Nduguru, Agnes Jonathan, Daniel Kandonga, Irene K Minja, Paschal Rugajo, Emmanuel Balandya and Julie Makani

**Writing – Review & Editing:** Upendo Masamu, Raphael Z Sangeda, Josephine Mgaya, Siana Nkya, Beatrice Octavian, Frank R Mtiiye, Joyce Nduguru, Agnes Jonathan, Daniel Kandonga, Irene K Minja, Paschal Rugajo, Emmanuel Balandya and Julie Makani.

